# The Prevalence of Non-Adherence in Patients with Resistant Hypertension: a Systematic Review and Meta-Analysis

**DOI:** 10.1101/2020.08.14.20175125

**Authors:** Gabrielle Bourque, Julius Vladimir Ilin, Marcel Ruzicka, Alexandra Davis, Gregory Hundemer, Swapnil Hiremath

**Affiliations:** The Ottawa Hospital and the University of Ottawa, 501 Smyth Road Ottawa, ON, Canada K1H8L6; The University of Ottawa, 451 Smyth Road Ottawa, ON, Canada K1H8L1; The Ottawa Hospital and the University of Ottawa, 1967 Riverside Drive Ottawa, ON, Canada K1H7W9; The Ottawa Hospital, 501 Smyth Road Ottawa, ON, Canada K1H8L6

## Abstract

**Background:** Resistant hypertension is quite prevalent and a risk factor for cardiovascular events. Patients with suspected resistant hypertension undergo more screening intensity for secondary hypertension, despite some of them being non-adherent to prescribed pharmacotherapy. The prevalence of non-adherence in this setting varies from about 5 to 80% in the published literature. Apart from the wide range, the relation between method of assessment and prevalence is not well established. Our objective was to establish the overall prevalence of non-adherence in the apparent treatment resistant hypertension population, explore causes of heterogeneity, and evaluate the effect of the method of assessment on the estimate of non-adherence.

**Methods:** We performed a systematic review and meta-analysis. MEDLINE, EMBASE Classic+EMBASE, Cochrane, CINAHL, and Web of Science were searched for relevant articles. Details about the method of adherence assessment were extracted from each included article and grouped into direct and indirect. Pooled analysis was performed using the random effects model and heterogeneity was explored with metaregression and subgroup analyses.

**Results:** The literature search yielded 1428 studies, of which 36 were included. The pooled prevalence of non-adherence was 35% (95% confidence interval 25 – 46 %). For indirect methods of adherence assessment, it was 25% (95% CI 15 – 39 %), whereas for direct methods of assessment, it was 44% (95% CI 32 – 57 %). Metaregression suggested gender, age, and time of publication as potential factors contributing to the heterogeneity.

**Conclusions:** Non-adherence to pharmacotherapy is quite common in resistant hypertension, with the prevalence varying with the methods of assessment.

**Brief Summary:** Resistant hypertension is known to be a risk factor for cardiovascular events. These patients also undergo higher screening intensity for secondary hypertension. However, not all patients with apparent treatment resistant hypertension have true resistant hypertension, with some of them being non-adherent to prescribed pharmacotherapy. This systematic review aims to establish the overall prevalence of non-adherence in the apparent treatment resistant hypertension population and assess the relative contributions of non-adherence assessed with direct and indirect measures.

## INTRODUCTION

Resistant hypertension (RH) is defined as blood pressure (BP) remaining above target despite the concurrent use of three or more antihypertensive agents of different classes, with one of the classes preferably being a diuretic and all of the medications being prescribed at optimal doses^1,2,3^. RH is common, with an estimated prevalence of about 10 to 30 %, and known to be a risk factor for cardiovascular events^4^. Furthermore, patients with RH are often part of high-risk groups with multiple cardiovascular comorbidities as well as vulnerable or disadvantaged populations. These patients undergo higher screening intensity for secondary hypertension. However, not all patients with apparent treatment RH have true RH. Apart from white coat hypertension and suboptimal prescribed drug doses and combinations, some individuals with apparent treatment RH may be non-adherent to the prescribed pharmacotherapy^5,6^.

There are several ways of assessing medication non-adherence, which can be broadly divided into indirect and direct methods. Indirect methods include questionnaires, self-reports, pill counts, rates of prescription refills, medication event monitoring systems (MEMS), and patient diaries. Direct methods include BP response to directly observed therapy (DOT) and measurement of the levels of BP-lowering drugs in physiologic fluids such as blood and urine^5^. The long term prevalence of non-adherence in chronic diseases is reported at about 50%^7^. However, this varied from 3 to 86% in individual studies in apparent treatment RH patients from a recent systematic review^8^. Interestingly, the pooled prevalence in this review varied based on the method of adherence measurement, from a low of 13% (similar estimate from self-report and physician interview) and 19% (prescription refill) to a high of 45% (DOT) and 49% (physical test, i.e. blood or urine assay). Though they were not grouped in this fashion, the former are indirect measures and the latter are direct measures. Apart from potentially different accuracy, these methods may capture different facets of non-adherence which might require different management strategies. As an example, pill counts and pharmacy refill data might identify occasional forgetfulness and/or carelessness, which can sometimes be related to an inability to follow instructions, either because of cognitive or physical limitations^5,7^. It can be managed using reminders, pill packs, and other interventions. In other settings, a patient may choose to alter the prescribed medication regimen, either by discontinuing medications entirely, skipping doses, or modifying doses or dosing intervals, however still continuing to refill prescriptions^6^. Underlying health beliefs and certain demographic factors and comorbid conditions may be associated with this, which would evade detection by indirect measures^9,10^. It requires more intensive measures (such as therapeutic drug monitoring (TDM) or DOT) to diagnose^5,6,11^. Interventions to address this form of non-adherence are also not well studied.

In the present study, we report on the overall prevalence of non-adherence in the apparent treatment RH population and what are the relative contributions of non-adherence based on different methods of assessment, with an emphasis on attempting to explain the heterogeneity of the data.

## METHODS

This systematic review and meta-analysis has been conducted and is reported in accordance with the Preferred Reporting Items for Systematic Reviews and Meta-Analyses (PRISMA) statement^12^. It is registered with the International Prospective Register of Systematic Reviews database (registration number: CRD42020137944). The study protocol has also been published separately^13^.

### Search Strategy

The databases MEDLINE (Ovid Interface, 1946 through April 02, 2019), EMBASE Classic+EMBASE (1947 through April 02, 2019), Cochrane (Cochrane Database of Systematic Reviews and Cochrane Central Register of Controlled Trials), Cumulative Index of Nursing and Allied Health Literature (CINAHL), and Web of Science were searched for relevant articles for this systematic review and meta-analysis. The search was designed by an information specialist (AD) and the search terms were adapted for the different databases. The MEDLINE search strategy is included in the supplementary appendix, table 1. Additional relevant records were identified through review of references of selected articles and from suggestions from experts in the field.

### Inclusion Criteria

We included observational studies, including cross sectional, retrospective, and prospective studies, as well as randomized controlled trials (RCTs) done on adult human participants aged 18 years or older with a diagnosis of RH, either uncontrolled on three or more drugs, or controlled with four or more drugs^1,2,3^. We only included studies where adherence to blood pressure lowering medications was measured using a test of adherence, either direct (such as TDM or DOT) or indirect (for example pill counts or pharmacy refill data). Only studies published in the English language were included. Studies published in other languages were included if a full-text version was available in English.

### Study Selection

Titles and abstracts of studies identified through the various database searches were uploaded to Covidence, an Internet based software program that facilitates collaboration among reviewers during the study selection process^14^. Two reviewers (GB and JVI) independently screened the titles and abstracts retrieved after the literature search to evaluate whether they met the pre-defined inclusion criteria. Conflicts between reviewers were resolved through discussion between the two reviewers until a consensus was reached. Full-text articles for the studies meeting inclusion criteria were retrieved and screened by the same two reviewers in order to select studies to be included in the systematic review. The reasons for excluding trials were recorded, both after title and abstract screening and after full-text screening. Reviewers were not blinded to the authors or journals when screening articles.

### Data Extraction

We extracted key elements of study design, definitions of RH used, demographic characteristics, comorbidities, and data on adherence from the included studies. For adherence, details about the method of assessment were extracted and grouped into direct (eg urine or plasma assay or TDM, DOT) and indirect (eg pill counts, questionnaires, adherence scales). For studies that reported more than one method of assessment, the highest value was used for the pooled analysis. Additionally, for studies that reported both direct and indirect methods, these were compared in a separate meta-analysis. All data was extracted in duplicate by two reviewers (GB and JVI).

### Quality and Risk of Bias Assessment

The study quality and the presence of potential bias within individual studies included in this systematic review were evaluated at both the outcome and study levels. The methodological quality of eligible full-text articles was independently assessed by two reviewers (GB and JVI) using the Newcastle-Ottawa scale^15^. The Newcastle-Ottawa scale includes the following domains: selection, comparability (not applicable to the studies included in this systematic review and meta-analysis), exposure (not applicable to the studies included in this systematic review and meta-analysis), and outcome. Disagreements were resolved through discussion until consensus was reached.

### Data Synthesis and Statistical Analysis

A pooled estimate of the prevalence of non-adherence was generated. The summary prevalence was estimated using the random effects modeling as described by DerSimonian-Laird^16^. We chose the random effects method because of its conservative summary estimate and because it incorporates between- and within-study variance. To assess heterogeneity of the event frequencies across studies, we used the Cochran’s Q-statistic test and the *I*^2^ statistic. All analyses were conducted using the Comprehensive Metaanalysis V2 software (Version 2.2, Biostat, Englewood NJ). Subgroup analyses were used to explore possible sources of heterogeneity, based on type of test used to measure adherence (grouped as direct versus indirect), study design, and definition of RH. We conducted univariate metaregression to assess moderator variables which are continuous in nature. The subgroup analyses and metaregression were also assessed as a method of resolving any statistical heterogeneity. Sensitivity analyses were conducted by excluding one study at a time and observing change in pooled estimate (with a > 10% change being considered significant). Publication bias was assessed by visual examination of the funnel plot and the Duval and Tweedie’s trim and fill method^17,18^.

## RESULTS

### Literature Search

The figure 1 shows a flow diagram of the systematic review process. The initial database search identified 2770 articles and 13 additional records were identified through other sources. After removal of duplicates, 1428 titles and abstracts were screened. 1218 articles were excluded at this stage and therefore 210 full-text articles were assessed for eligibility. 174 of these were excluded, with the most common reasons for exclusion being absence of RH as defined above and ineligible study design, such as case reports, reviews, or editorials. Therefore, 36 full-text articles were eligible for inclusion in this systematic review and meta-analysis. A summary of the included articles is included in Legend for **Figures and Tables**

**Figure 1:**
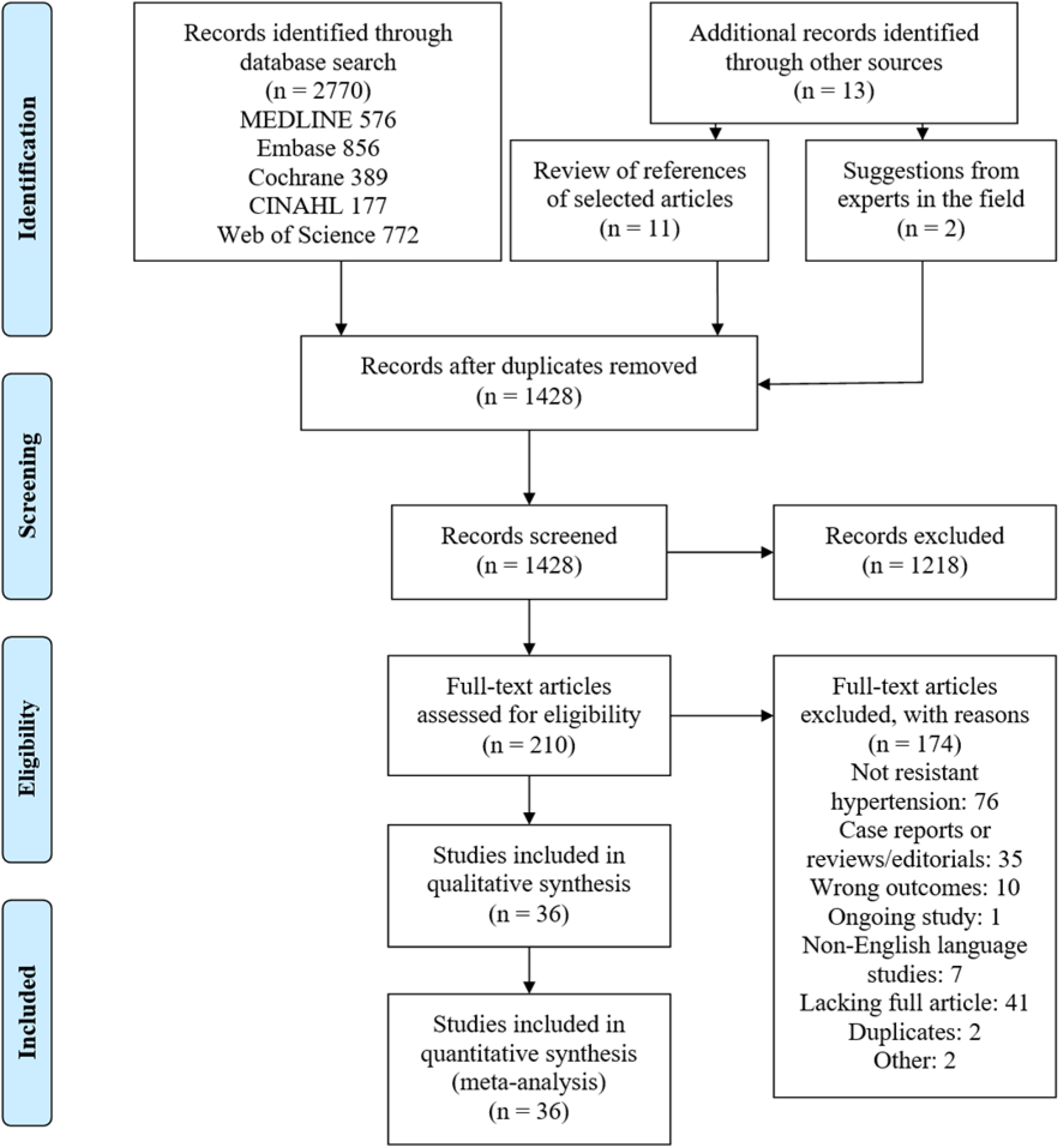
Flow Diagram for Literature Search and Selection

Table 1 Study Characteristics

Table 2 Population Characteristics

Figure 1 Flow Diagram for Literature Search and Selection

Figure 2 Forest Plot

### Supplementary Appendix

Supplementary Table 1 MEDLINE Search Strategy – Ovid Interface

Supplementary Table 2 Quality Assessment

Supplementary Figure 1 Regression of Mean Age on Logit Event Rate

Supplementary Figure 2 Regression of Proportion of Men on Logit Event Rate

Supplementary Figure 3 Regression of Publication date on Logit Event Rate

Supplementary Figure 4 Regression of Sample Size on Logit Event Rate

Supplementary Figure 5 Regression of Sample Size excluding one study on Logit Event Rate with Exclusion of One Large Study

***Table 1*** and Table 2, which describe the study and population characteristics.

**Table 1:**
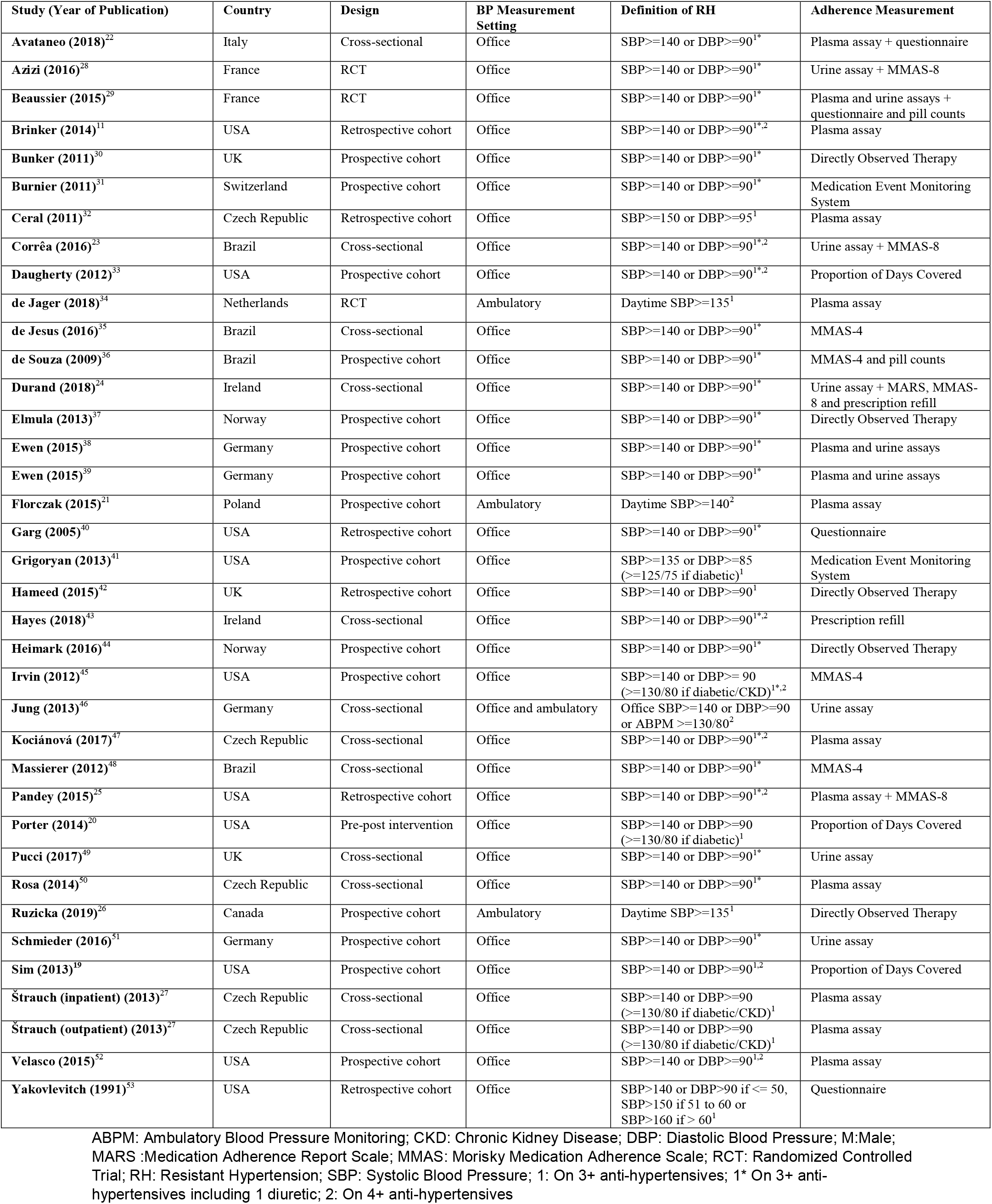
Study Characteristics

**Table 2:** Population Characteristics

| **Study (Year of Publication)** | **Sample Size** | **Sex (% M)** | **Mean Age (years ± standard deviation)** |
| --- | --- | --- | --- |
| **Avataneo (2018)**<sup>22</sup> | 50 | 58 | 56 ± 7 |
| **Azizi (2016)**<sup>28</sup> | 85 | NR | NR |
| **Beaussier (2015)**<sup>29</sup> | 164 | 55 | 55 ± 10 |
| **Brinker (2014)**<sup>11</sup> | 56 | NR | NR |
| **Bunker (2011)**<sup>30</sup> | 37 | 35 | 57 |
| **Burnier (2011)**<sup>31</sup> | 41 | 76 | 50.5 |
| **Ceral (2011)**<sup>32</sup> | 84 | 59.5 | 55 ± 11 |
| **Corrêa (2016)**<sup>23</sup> | 21 | 43 | NR |
| **Daugherty (2012)**<sup>33</sup> | 3550 | 49 | 60 |
| **de Jager (2018)**<sup>34</sup> | 98 | 42 | 63 ± 11 |
| **de Jesus (2016)**<sup>35</sup> | 96 | 43.8 | 53.9 ± 9.9 |
| **de Souza (2009)**<sup>36</sup> | 44 | 27.3 | 48.6 ± 9.0 |
| **Durand (2018)**<sup>24</sup> | 204 | 57.8 | 70 ± 11 |
| **Elmula (2013)**<sup>37</sup> | 18 | 89 | 54 |
| **Ewen (2015)**<sup>38</sup> | 27 | 60 | 61.9 ± 9.9 |
| **Ewen (2015)**<sup>39</sup> | 100 | 67 | 62.7 ± 10.6 |
| **Florczak (2015)**<sup>21</sup> | 36 | 64 | 52.5 ± 9.1 |
| **Garg (2005)**<sup>40</sup> | 141 | 45 | 57 ± 13 |
| **Grigoryan (2013)**<sup>41</sup> | 69 | NR | NR |
| **Hameed (2015)**<sup>42</sup> | 48 | 47.9 | 62 ± 11 |
| **Hayes (2018)**<sup>43</sup> | 646 | 53.7 | 71.1 ± 12.2 |
| **Heimark (2016)**<sup>44</sup> | 83 | 78 | 58.3 ± 10.0 |
| **Irvin (2012)**<sup>45</sup> | 2654 | 48.2 | 67.4 ± 8.7 |
| **Jung (2013)**<sup>46</sup> | 76 | 57.9 | 58 |
| **Kociánová (2017)**<sup>47</sup> | 106 | 56 | 56,8 |
| **Massierer (2012)**<sup>48</sup> | 106 | 27 | 57.3 ± 10.6 |
| **Pandey (2015)**<sup>25</sup> | 47 | 53 | 52.5 ± 2 |
| **Porter (2014)**<sup>20</sup> | 60 | 96.7 | 62 ± 6.18 |
| **Pucci (2017)**<sup>49</sup> | 131 | NR | NR |
| **Rosa (2014)**<sup>50</sup> | 72 | NR | 55.5 ± 10.5 |
| **Ruzicka (2019)**<sup>26</sup> | 48 | 66.7 | 62.1 ± 13.1 |
| **Schmieder (2016)**<sup>51</sup> | 79 | 72 | 60.4 ± 10 |
| **Sim (2013)**<sup>19</sup> | 60327 | 48 | 69 ± 11 |
| **Štrauch (inpatient) (2013)**<sup>27</sup> | 176 | 58,5 | 52 ± 11 |
| **Štrauch (outpatient) (2013)**<sup>27</sup> | 163 | 56,4 | 54 ± 12 |
| **Velasco (2015)**<sup>52</sup> | 78 | NR | NR |
| **Yakovlevitch (1991)**<sup>53</sup> | 91 | 42 | 57 ± 14 |

*M – Male; NR – Not Reported*

### Study Characteristics

36 studies were included in this systematic review. Of these, 21 were conducted in Europe, 11 in North America, and 4 in South America. The majority of the included studies were observational, with 15 prospective cohort studies, 11 cross-sectional studies, and 6 retrospective cohort studies. 3 of the included studies were RCTs and 1 was a pre-post intervention study.

The total number of participants included was 69,912. One study^19^ accounted for the vast majority of these. The median sample size was 83, with a range from 18 to 60,327. The proportion of patients with comorbidities, such as diabetes, chronic kidney disease (CKD), congestive heart failure (CHF), and cerebrovascular disease (CVD), was inconsistently reported in the individual studies.

### Definition of Resistant Hypertension

The definition of RH as well as the thresholds used to define non-adherence to prescribed pharmacotherapy varied between studies. 32 studies used office BP for defining RH and only 3 used ambulatory blood pressure monitoring (ABPM). One study used both methods. Antihypertensive regimens were not consistently reported and it often was not specified whether patients were on an optimized regimen. The most common threshold for diagnosis of RH was systolic blood pressure (SBP) ≥ 140 and/or diastolic blood pressure (DBP) ≥ 90. Patients with secondary forms of hypertension as well as white-coat hypertension were not consistently excluded, as is apparent from the low use of ABPM for diagnosis of RH.

### Assessment of Adherence

18 studies assessed non-adherence using direct methods, 12 with indirect methods, and 6 with both direct and indirect methods, either concurrently or sequentially (see Table 1). DOT was used in 5 studies, plasma and/or urine assays in 19, MEMS in 2, non-standardized questionnaires in 4, standardized questionnaires such as the Morisky Medication Adherence Scale (MMAS) and the Medication Adherence Report Scale (MARS) in 8, prescription refill in 2, Proportion of Days Covered (PDC) in 3, and pill counts in 2 (see Table 1).

### Prevalence of Non-Adherence

The pooled prevalence of non-adherence to antihypertensive pharmacotherapy was 35% (95% confidence interval [CI] 25 to 46%) as shown in figure 2. The prevalence of non-adherence varied across studies, with values ranging from 3.3%^20^ to 86.1%^21^, leading to a high degree of statistical heterogeneity (I^2^ = 99, p < 0.001).

**Figure 2:**
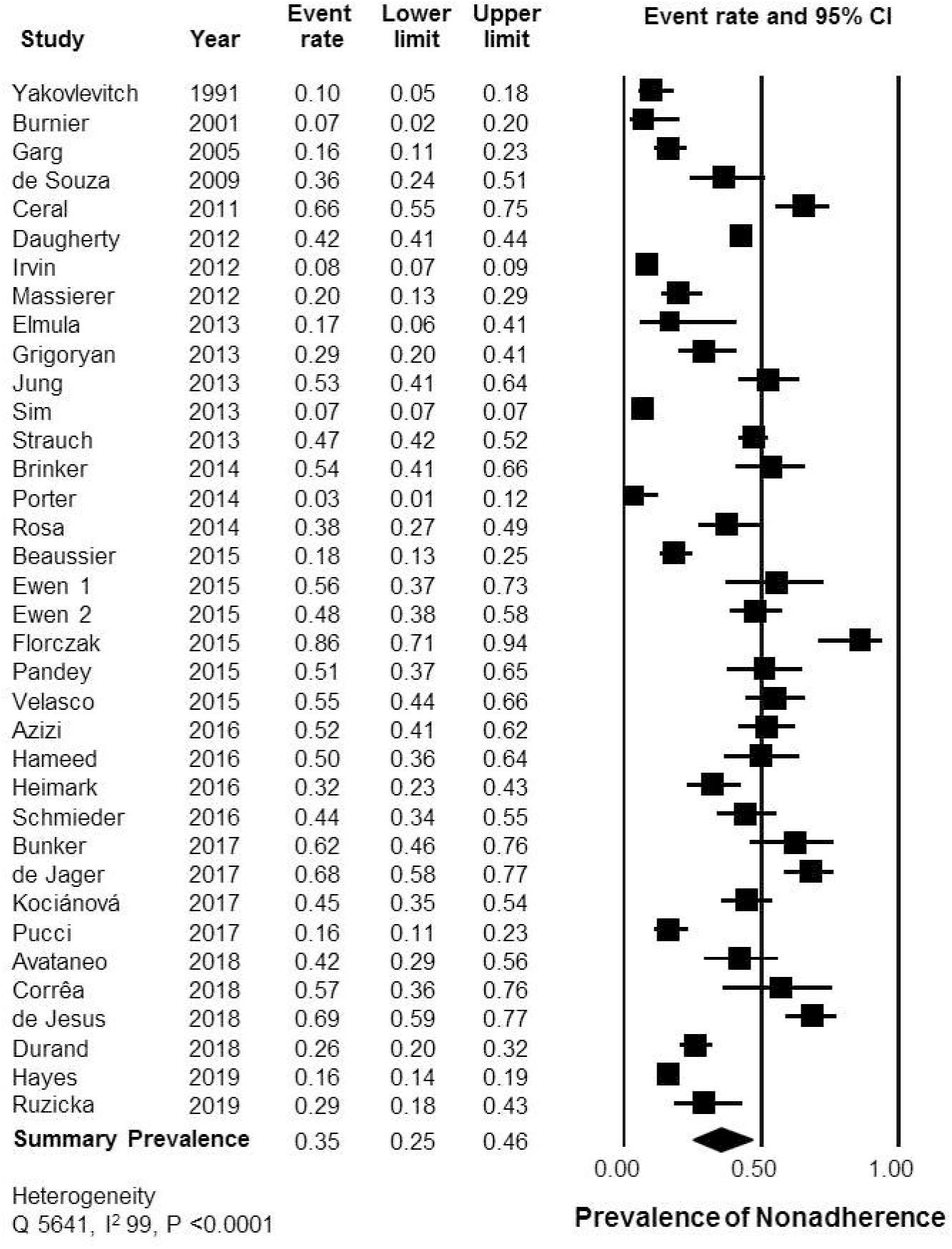
Forest Plot

### Subgroup Analyses and Metaregression

Using indirect methods of adherence assessment, the pooled prevalence of non-adherence was 25% (95% CI 15 – 39%, I^2^ = 99), whereas with direct methods of assessment, it was 44% (95% CI 32 – 57%, I^2^ = 89). The prevalence of non-adherence was higher in the subgroup of studies where ABPM was required for definition of RH (61%, 95% CI 27 – 86%, I^2^ = 89), compared to the subgroup of studies which defined RH based on office BP (32%, 95% CI 22 – 44%, I^2^ = 99). Lastly, there was no difference in the prevalence of adherence in retrospective studies (38%, 95% CI 16 – 66%, I^2^ = 94) or prospective studies (34%, 95% CI 24 – 47%, I^2^ = 99). Excluding one study did not change the overall pooled estimate by > 10% (range 34 – 36%, compared to 35% with all studies). The subgroup analyses did not resolve the heterogeneity, which remained high.

With univariate metaregression (supplementary appendix, figures 1 to 5), there was an association of higher prevalence of non-adherence with younger age (slope coefficient 0.20, p < 0.001), higher proportion of men (slope coefficient 0.08, p < 0.001), and with more recent studies (slope coefficient 0.04, p < 0.001). An association of larger sample size with lower prevalence of non-adherence was also seen (slope coefficient 0.003, p < 0.001), which was driven by one large study with a sample size of 60,327 and a non-adherence prevalence of 11%. Exclusion of this study resulted in the association no longer being significant (slope coefficient 0.0001, p = 0.40).

Six studies reported both direct and indirect methods of assessment^22,23,24,25,26,27^. In a pooled analysis, the odds of detecting non-adherence were higher with direct methods (summary odds ratio 1.95, 95% CI 1.31 to 1.95, p = 0.001), though with significant statistical heterogeneity (I^2^ = 98).

### Quality Assessment and Publication Bias

The overall quality of the studies was fair, with a median of 4 stars (range 3 to 5). A large proportion of studies rated high on the representativeness subsection of the selection domain of the Newcastle-Ottawa scale. However, they rated poorly on the subsection demonstration that the outcome of interest, in this case non-adherence to antihypertensive medications, was not present at start of study. They rated mostly high on the assessment of outcome, length of follow-up, and adequacy of follow-up subsections of the outcome domain of the scale (see supplementary appendix, table 2).

There was no evidence of publication bias from visual examination of the funnel plot and no additional studies were imputed using the Duval and Tweedie’s trim and fill method (see supplementary appendix, figure 6).

## DISCUSSION

In this systematic review, we report a high degree of non-adherence prevalence in patients with apparent treatment RH, at 35%. However, there was significant heterogeneity identified among the studies, with a range of non-adherence from 3% to 86%. Studies using direct methods reported a higher degree of non-adherence, at 44%, compared to indirect methods, at 25%. The study level metaregression reveals younger age, male sex, and more recent publication date as being associated with higher rates of non-adherence.

Though non-adherence has always been part of the evaluation for RH, the extent of non-adherence, approximately at 1 in 3 patients in this systematic review, is so high, that an accurate assessment of adherence is essential before the patient is labeled as RH. Instead of investigations for secondary hypertension, consuming valuable healthcare resources, more attention needs to be focused on assessment and management of non-adherence in these patients. Additionally, non-adherence in most clinical settings is assessed by patient questionnaire, pharmacy filling data, or pill counts. As we demonstrate, these indirect methods of adherence provide a lower estimate compared to direct methods of assessment of non-adherence, such as TDM or DOT. Unfortunately, both TDM and DOT are mostly research tools and not available or funded in routine clinical practice. We hope this systematic review elevates their status for more routine use, especially in the setting of RH, where the high prevalence of non-adherence makes these tools more valuable and possibly cost effective.

A previous systematic review^8^ also reports on a high degree of non-adherence in this population, though with 24 studies, compared to 36 in the present study. Additionally, we believe grouping assessment methods into direct and indirect methods provides a more accurate picture of the non-adherence phenotype. Additionally, in the present study, the exploration of heterogeneity provides more insight into the fact that patients of a younger age and men might have a higher non-adherence phenotype. We also report the trend for higher rates of non-adherence in the recently published studies. It is not quite clear whether this is because of ascertainment bias or more accurate assessment of non-adherence. Regardless, pill counts and pharmacy filling data may be necessary, but are far from sufficient for assessment of adherence.

The dichotomy of direct and indirect assessment of adherence does not necessarily mean that one is more accurate than the other. Indirect assessment of non-adherence with pill counts and pharmacy filling data may provide more insight into the occasional forgetfulness or missed doses in a patient who is otherwise engaged with the medication regimen. Direct methods of non-adherence provide pharmacokinetic and pharmacodynamic data, however in a cross-sectional manner. We would posit that these two broad techniques are complementary in the complete diagnosis of non-adherence.

The assessment of non-adherence is merely the first step in adequate blood pressure control. Management of non-adherence requires further exploration of the reasons of non-adherence and taking steps accordingly. Much of the focus has been on patient aids, such as reminders and smart pill bottles, and these would be useful for the unintentional non-adherence phenotype. For patients who are otherwise disengaged in the medication and therapy plan, further exploration of their motives and beliefs is necessary. The diagnosis of non-adherence is but the initial step in this process.

Possible limitations of this study include the finding of severe heterogeneity, which has not been completely resolved by the analytic plan, the limitations of the literature search), and the potential lack of granular data in the published studies for a patient-level meta-analysis. However, there was no publication bias seen in the analysis and an information specialist was used for the literature search, attenuating these issues.

## CONCLUSIONS

In patients with apparent treatment RH, standard indirect measures of assessing adherence can miss a substantial proportion of patients who are non-adherent to prescribed pharmacotherapy.

This systematic review and meta-analysis also provides a basis for future research on strategies to better address the different factors that contribute to medication non-adherence in this setting.

## Data Availability

We only use publicly available data

## ACKNOWLEDGEMENTS

SH, GH, and MR receive research salary support from the Department of Medicine, University of Ottawa.

## LIST OF ABBREVIATIONS

ABPM: Ambulatory Blood Pressure Monitoring
BP: Blood Pressure
CHF: Congestive Heart Failure
CI: Confidence Interval
CINHAL: Cumulative Index of Nursing and Allied Health Literature
CKD: Chronic Kidney Disease
CVD: Cerebrovascular Disease
DBP: Diastolic Blood Pressure
DOT: Directly Observed Therapy
HR: Heart Rate
MARS: Medication Adherence Report Scale
MEMS: Medication Event Monitoring System
MMAS: Morisky Medication Adherence Scale
PDC: Proportion of Days Covered
PRISMA: Preferred Reporting Items for Systematic Reviews and Meta-Analyses
RCTs: Randomized Controlled Trials
RH: Resistant Hypertension
SBP: Systolic Blood Pressure
TDM: Therapeutic Drug Monitoring

## REFERENCES

1. Calhoun DA, Jones D, et al. Resistant Hypertension: Diagnosis, Evaluation, and Treatment. A Scientific Statement from the American Heart Association Professional Education Committee of the Council for High Blood Pressure Research. Hypertension 2008; 51: 1403–1419.

2. Mancia G, Fagard R, Narkiewicz K, et al. 2013 ESH/ESC Guidelines for the Management of Arterial Hypertension: the Task Force for the Management of Arterial Hypertension of the European Society of Hypertension (ESH) and of the European Society of Cardiology (ESC). Blood Press 2013; 22: 193–278.

3. Hiremath S, Sapir-Pichhadze R, Nakhla M, et al. Hypertension Canada’s 2020 Evidence Review and Guidelines for the Management of Resistant Hypertension. Can J Cardiol 2020; 36(5): 625–634.

4. Sinnott SJ, Smeeth L, Williamson E, Douglas IJ. Trends for Prevalence and Incidence of Resistant Hypertension: Population Based Cohort Study in the UK 1995–2015. BMJ 2017; 358: j3984.

5. Ruzicka M, Hiremath S. Can Drugs Work in Patients who do Not Take Them? The Problem of Non-Adherence in Resistant Hypertension. Curr Hypertens Rep 2015 Sep; 17(9): 579.

6. Ruzicka M, McCormick B, Leenen FH, Froeschl M, Hiremath S. Adherence to Blood Pressure-Lowering Drugs and Resistant Hypertension: Should Trial of Direct Observation Therapy be Part of Preassessment for Renal Denervation? Can J Cardiol 2013; 29(12): 1741 e1–3.

7. Burnier M, Wuerzner G, Struijker-Boudier H, Urquhart J. Measuring, Analyzing, and Managing Drug Adherence in Resistant Hypertension. Hypertension 2013; 62(2): 218–25.

8. Durand H, Hayes P, Morrissey EC, et al. Medication Adherence among Patients with Apparent Treatment-Resistant Hypertension: Systematic Review and Meta-Analysis. J Hypertens 2017; 35(12): 2346–57.

9. Griva K, Neo HLM, Vathsala A. Unintentional and Intentional Non-Adherence to Immunosuppressive Medications in Renal Transplant Recipients. Int J Clin Pharm 2018; 40(5): 1234–1241.

10. Mukhtar O, Weinman J, Jackson SH. Intentional Non-Adherence to Medications by Older Adults. Drugs Aging 2014; 31(3): 149–157.

11. Brinker S, Pandey A, Ayers C, et al. Therapeutic Drug Monitoring Facilitates Blood Pressure Control in Resistant Hypertension. J Am Coll Cardiol 2014; 63(8): 834–835.

12. Moher D, Liberati A, Tetzlaff J, Altman DG; PRISMA Group. Preferred Reporting Items for Systematic Reviews and Meta-Analyses: the PRISMA Statement. Int J Surg 2010; 8(5):336–341.

13. Bourque G, Ilin JV, Ruzicka M, Davis A(S), Hiremath S. The Prevalence of Non-Adherence in Patients with Resistant Hypertension: a Systematic Review Protocol. Can J Kidney Health Dis 2019; 6:1–7.

14. Covidence – Better Systematic Review Management. World-Class Systematic Review Management. Available at: https://www.covidence.org/home. Accessed on October 23, 2019.

15. Wells GA, Shea B, O’Connell D, et al. The Newcastle-Ottawa Scale (NOS) for Assessing the Quality of Nonrandomised Studies in Meta-Analyses. Available at: www.ohri.ca/programs/clinical_epidemiology/oxford.asp. Accessed on July 9, 2019.

16. DerSimonian R, Laird N. Meta-Analysis in Clinical Trials. Control Clin Trials 1986; 7(3): 177–188.

17. Egger M, Davey Smith G, Schneider M, et al. Bias in Meta-Analysis Detected by a Simple, Graphical Test. BMJ 1997; 315(7109): 629–634.

18. Duval S, Tweedie R. Trim and Fill: A Simple Funnel-Plot-Based Method of Testing and Adjusting for Publication Bias in Meta-Analysis. Biometrics 2000; 56(2): 455–463.

19. Sim JJ, Bhandari SK, Shi J, et al. Characteristics of Resistant Hypertension in a Large Ethnically Diverse Hypertension Population of an Integrated Health System. Mayo Clin Proc 2013; 88(10): 1099–1107.

20. Porter AK, Taylor SR, Yabut AH, Al-Achi A. Impact of a Pill Box Clinic to Improve Systolic Blood Pressure in Veterans with Uncontrolled Hypertension Taking 3 or More Antihypertensive Medications. J Manag Care Spec Pharm 2014; 20(9): 905–911.

21. Florczak E, Tokarcyk B, Warchol-Celińska E, et al. Assessment of Adherence to Treatment in Patients with Resistant Hypertension using Toxicological Serum Analysis. A Subgroup Evaluation of the RESIST-POL Study. Pol Arch Med Wewn 2015; 125(1–2): 65–72.

22. Avataneo V, de Nicolò A, Rabbia F, et al. Therapeutic Drug Monitoring-Guided Definition of Adherence Profiles in Resistant Hypertension and Identification of Predictors of Poor Adherence. Br J Clin Pharmacol 2018; 84: 2535–2543.

23. Corrêa NB, de Faria AP, Ritter AM, et al. A Practical Approach for Measurement of Antihypertensive Medication Adherence in Patients with Resistant Hypertension. J Am Soc Hypertens 2016; 10(6): 510–516.

24. Durand H, Hayes P, Harhen B, et al. Medication Adherence for Resistant Hypertension: Assessing Theoretical Predictors of Adherence using Direct and Indirect Adherence Measures. Br J Health Psychol 2018; 23(4): 949–966.

25. Pandey A, Raza F, Velasco A, et al. Comparison of Morisky Medication Adherence Scale with Therapeutic Drug Monitoring in Apparent Treatment-Resistant Hypertension. J Am Soc Hypertens 2015; 9(6): 420–426.

26. Ruzicka M, Leenen FHH, Ramsay T, et al. Use of Directly Observed Therapy to Assess Treatment Adherence in Patients with Apparent Treatment-Resistant Hypertension. JAMA Intern Med 2019; 1455.

27. Štrauch B, Petrák O, Zelinka T, et al. Precise Assessment of Noncompliance with the Antihypertensive Therapy in Patients with Resistant Hypertension using Toxicological Serum Analysis. J Hypertens 2013; 31(12): 2455–2461.

28. Azizi M, Pereira H, Hamdidouch I, et al. Adherence to Antihypertensive Treatment and the Blood Pressure-Lowering Effects of Renal Denervation in the Renal Denervation for Hypertension (DENERHTN) Trial. Circulation 2016; 134(12): 847–857.

29. Beaussier H, Boutouyrie P, Bobrie G, et al. True Antihypertensive Efficacy of Sequential Nephron Blockade in Patients with Resistant Hypertension and Confirmed Medication Adherence. J Hypertens 2015; 33(12): 2526–2533.

30. Bunker J, Callister W, Chang CL, Sever PS. How Common is True Resistant Hypertension? J Hum Hypertens 2011; 25(2): 137–140.

31. Burnier M, Schneider MP, Chioléro A, Stubi CLF, Brunner HR. Electronic Compliance Monitoring in Resistant Hypertension: the Basis for Rational Therapeutic Decisions. J Hypertens 2001; 19(2): 335–341.

32. Ceral J, Habrdova V, Vorisek V, Bima M, Pelouch R, Solar M. Difficult-to-Control Arterial Hypertension or Uncooperative Patients? The Assessment of Serum Antihypertensive Drug Levels to Differentiate Non-Responsiveness from Non-Adherence to Recommended Therapy. Hypertens Res 2011; 34(1): 87–90.

33. Daugherty SL, Powers JD, Magid DJ, et al. The Association between Medication Adherence and Treatment Intensification with Blood Pressure Control in Resistant Hypertension. Hypertension 2012; 60(2): 303–309.

34. de Jager RL, van Maarseveen EM, Bots ML, Blankestijn PJ, and on behalf of the SYMPATHY investigators. Medication adherence in patients with apparent resistant hypertension: findings from the SYMPATHY trial. Br J Clin Pharmacol 2018; 84(1): 18–24.

35. de Jesus NS, Nogueira AD, Pachu CO, Luiz RR, de Oliveira GMM. Blood Pressure Treatment Adherence and Control after Participation in the ReHOT. Arq Bras Cardiol 2016; 107(5): 437–445.

36. de Souza WA, Sabha M, de Faveri Favero F, Bergsten-Mendes G, Yugar-Toledo JC, Moreno Jr H. Intensive Monitoring of Adherence to Treatment Helps to Identify ‘‘True’’ Resistant Hypertension. J Clin Hypertens (Greenwich) 2009; 11(4): 183–191.

37. Elmula FEMF, Hoffmann P, Fossum E, et al. Renal Sympathetic Denervation in Patients with Treatment-Resistant Hypertension after Witnessed Intake of Medication before Qualifying Ambulatory Blood Pressure. Hypertension 2013; 62(3): 526–532.

38. Ewen S, Cremers B, Meyer MR, et al. Blood Pressure Changes after Catheter-Based Renal Denervation are Related to Reductions in Total Peripheral Resistance. J Hypertension 2015; 33(12): 2519–2525.

39. Ewen S, Meyer MR, Cremers B, et al. Blood Pressure Reductions following Catheter-Based Renal Denervation are not Related to Improvements in Adherence to Antihypertensive Drugs Measured by Urine/Plasma Toxicological Analysis. Clin Res Cardiol 2015; 104(12): 1097–1105.

40. Garg JP, Elliott WJ, Folker A, Izhar M, Black HR. Resistant Hypertension Revisited: a Comparison of Two University-Based Cohorts. Am J Hypertens 2005; 18(5 Pt 1): 619–626.

41. Grigoryan L, Pavlik VN, Hyman DJ. Characteristics, Drug Combinations and Dosages of Primary Care Patients with Uncontrolled Ambulatory Blood Pressure and High Medication Adherence. J Am Soc Hypertens 2013; 7(6): 471–476.

42. Hameed MA, Tebbit L, Jacques N, Thomas M, Dasgupta I. Non-Adherence to Antihypertensive Medication is Very Common among Resistant Hypertensives: Results of a Directly Observed Therapy Clinic. J Hum Hypertens 2016; 30(2): 83–89.

43. Hayes P, Casey M, Glynn LG, et al. Prevalence of Treatment-Resistant Hypertension after Considering Pseudo-Resistance and Morbidity: a Cross-Sectional Study in Irish Primary Care. Br J Gen Pract 2018; 68(671): e394–e400.

44. Heimark S, Eskås PA, Mariampillai JE, Larstorp ACK, Høieggen, Elmula FEMF. Tertiary Work-Up of Apparent Treatment-Resistant Hypertension. Blood Press 2016; 25(5): 312–318.

45. Irvin MR, Shimbo D, Mann DM, et al. Prevalence and Correlates of Low Medication Adherence in Apparent Treatment Resistant Hypertension. J Clin Hypertens (Greenwich) 2012; 14(10): 694–700.

46. Jung O, Gechter JL, Wunder C, et al. Resistant Hypertension? Assessment of Adherence by Toxicological Urine Analysis. J Hypertens 2013; 31(4): 766–774.

47. Kociánová E, Václavík J, Tomková J, et al. Heart Rate is a Useful Marker of Adherence to Beta-Blocker Treatment in Hypertension. Blood Press 2017; 26(5): 311–318.

48. Massierer D, Oliveira AC, Steinhorst AM, et al. Prevalence of Resistant Hypertension in Non-Elderly Adults: Prospective Study in a Clinical Setting. Arq Bras Cardiol 2012; 99(1): 630–635.

49. Pucci M, Martin U. Detecting Non-Adherence by Urine Analysis in Patients with Uncontrolled Hypertension: Rates, Reasons and Reactions. J Hum Hypertens 2017; 31(4): 253–257.

50. Rosa J, Zelinka T, Petrák O, et al. Importance of Thorough Investigation of Resistant Hypertension before Renal Denervation: Should Compliance to Treatment be Evaluated Systematically? J Hum Hypertens 2014; 28(11): 684–688.

51. Schmieder RE, Ott C, Schmid A, et al. Adherence to Antihypertensive Medication in Treatment-Resistant Hypertension Undergoing Renal Denervation. J Am Heart Assoc 2016; 5(2).

52. Velasco A, Chung O, Raza F, et al. Cost Effectiveness of Therapeutic Drug Monitoring in Diagnosing Primary Aldosteronism in Patients with Resistant Hypertension. J Clin Hypertens (Greenwich) 2015; 17(9): 713–719.

53. Yakovlevitch M, Black HR. Resistant Hypertension in a Tertiary Care Clinic. Arch Intern Med 1991; 151(9): 1786–1792.

